# Ambler class C-type β-lactamases in *Enterobacter* spp. and *Klebsiella aerogenes* in the Netherlands, 2012-2023

**DOI:** 10.1101/2025.02.19.25322487

**Authors:** Elke van Gorp, Simon Lansu, Cornelia C.H. Wielders, Angela de Haan, Gert-Jan Godeke, Sandra Witteveen, Jeroen Bos, Fabian Landman, Maureen Ouw, Herman F. Wunderink, Maurits P.A. van Meer, Wil van der Zwet, Daan W. Notermans, Antoni P.A. Hendrickx the Dutch CPE Surveillance Study Group

## Abstract

We investigated the genomic epidemiology of Ambler class C (AmpC-type) β-lactamases in *Enterobacter* spp. and *Klebsiella aerogenes* in the national carbapenemase-producing Enterobacterales (CPE) surveillance of the Netherlands between 2012 and 2023. A total of 399 *E. cloacae* complex and *K. aerogenes* isolates from 399 patients were analyzed using whole-genome sequencing to assess genetic relatedness, resistance gene profiles, plasmid replicons, and the genomic location of AmpC-genes, respectively. Of the 399 patients, 217 were male (54%), and the median age was 67 years. Carbapenemase production was assessed using the carbapenem inactivation method (CIM) and CarbaNP-test. Considerable proportions of *Enterobacter* spp. (32%) and *K. aerogenes* (52%) isolates produced carbapenemase, without detectable major carbapenemase genes (IMP, KPC, NDM, OXA-48-like, VIM), a phenotype termed CIM+Carba-. These isolates were mostly (82%) susceptible (EUCAST ≤2mg/L) to meropenem. The majority of CIM+Carba+ isolates with major carbapenemase genes were gained from pre-emptive screening, while CIM+Carba-isolates were mainly taken for diagnostic purposes. Genomic analysis identified 18 genogroups, with *E. kobei*, *E. roggenkampii*, *E. ludwigii,* and *K. aerogenes* showing the CIM+Carba-phenotype, correlating with chromosome-encoded AmpC-type β-lactamases like *bla*_ACT-28_, *bla*_ACT-52,_ *bla*_MIR-3_, *bla*_MIR-11_ or *ampC* of which the majority (63%) yielded a positive CarbaNP. These CIM+Carba-isolates carried only few plasmids, and there was limited nosocomial spread. CIM+Carba-*E. kobei* carrying *bla*_ACT-28_ overproduced ACT-28 protein in the CIM. Overall, the *Enterobacter* and *K. aerogenes* population in the Netherlands is genetically diverse, with most isolates carrying species-specific AmpC-type β-lactamases with putative carbapenemase activity and represent a low-risk for public health.

**Importance:** CPE represents an important healthcare problem worldwide. This study highlights the diverse genetic *Enterobacter* spp. population in the Dutch CPE surveillance with *E. hormaechei* subsp. *steigerwaltii* as the most common carbapenemase-producing (CIM+Carba+) species. However, a significant proportion of *E. kobei*, *E. roggenkampii*, *E. ludwigii,* and *K. aerogenes* obtained in the Netherlands carry chromosomal AmpC-type β-lactamases with putative carbapenemase activity in the absence of major carbapenemases (CIM+Carba-), were mostly susceptible for meropenem, and showed limited nosocomial spread. We recommend whole-genome sequencing for accurate *Enterobacter*/*K. aerogenes* species classification, and AmpC-type β-lactamase gene identification. Despite limited carbapenem resistance and dissemination, proper infection control measures are necessary. The work outlined here underscores the importance of distinguishing *E. kobei*, *E. roggenkampii*, *E. ludwigii,* and *K. aerogenes* isolates and its AmpC-type β-lactamases from true CPE by whole-genome sequencing to avoid misclassification and unnecessary infection prevention and public health interventions.

## Introduction

Carbapenemase-producing Enterobacterales (CPE) represent a global problem due to limited options for antibiotic treatment, increased healthcare costs, and increased morbidity and mortality, particularly among hospitalized patients (1). In 2024, the World Health Organization prioritized third-generation cephalosporin and carbapenem-resistant Enterobacterales as a critical-priority pathogen for research and development of new antibiotics (2). Carbapenemases are a subgroup of the β-lactamases which can hydrolyse most β-lactams, including carbapenems (3). β-lactamase enzymes can be divided into four Ambler classes, class A to D, based on their amino acid sequence (3-5). Ambler class A enzymes group the penicillinases, extended-spectrum β-lactamases (ESBLs) and serine carbapenemases (e.g. KPC), class B encompasses the metallo-β-lactamases (e.g. IMP, NDM, VIM), class C belongs to the cephalosporinases (e.g. AmpC) and class D entails the oxacillinases (e.g. OXA-48-like) (4). To date, IMP, KPC, NDM, OXA-48, and VIM are clinically the most important and best characterized carbapenemases that have spread globally among Enterobacterales (6, 7).

*Enterobacter cloacae* complex, belonging to the order of Enterobacterales, is a group of Gram-negative opportunistic pathogens with large ecological distribution, and is considered to be part of the ESKAPE pathogens causing hospital infections (8). *E*. *cloacae* complex ranked third (13%) in frequency among Enterobacterales species sent to the National Institute for Public Health and the Environment (RIVM) for CPE surveillance in the Netherlands between 2014-2018 (9). The *E. cloacae* complex comprises *E. asburiae*, *E. cloacae*, *E. hormaechei*, *E. kobei*, *E. ludwigii*, *E. mori*, and *E. nimipressuralis,* and acquires a wide range of antimicrobial resistance (AMR) genes and plasmids (8, 10). Ambler class C enzymes were originally regarded as chromosomally localized enzymes of *Enterobacter*, but are found on plasmids as well (11). Even though Ambler class C β-lactamase genes are considered to have minimal activity against carbapenems, some AmpC genes, such as *bla*_ACT-28_, have been shown to exhibit weak carbapenemase activity *in vitro* (12-14).

The major objective of this study was to analyze the genomic epidemiology of Ambler class C-type β-lactamase-carrying *Enterobacter* spp. and *K. aerogenes* which had a positive test result for the carbapenem inactivation method (CIM), while lacking IMP-, KPC-, NDM-, OXA-48-like-, and VIM-type enzymes obtained in the Dutch national CPE surveillance from 2012-2023.

## Materials and Methods

### Bacterial isolates

The RIVM conducts national surveillance on CPE. Medical microbiology laboratories (MML) in the Netherlands are requested to submit suspected CPE isolates to the RIVM for characterization. Submission criteria are genotypic evidence or phenotypic carbapenemase production, minimum inhibitory concentration (MIC) of >0.25 mg/L for meropenem and/or >1 mg/L for imipenem (9, 15). Only the first submitted Enterobacterales isolate per-person-per-species-per-carbapenemase-allele, submitted during the period 2012-2023, were included in this study. Extensive analysis was performed on a subset of *E. cloacae* complex isolates for which next-generation sequencing (NGS) data was available. *K. aerogenes* isolates, previously classified as *E. aerogenes*, were also included in this subset.

### Antimicrobial susceptibility testing and phenotypic analysis of bacterial isolates

For all isolates, the MIC for meropenem was determined by Etest (BioMérieux Inc., Marcy l’Etoile, France) and the carbapenemase production was phenotypically assessed by the CIM (16). On a subset of isolates the CarbaNP test (BioMérieux Inc., Marcy l’Etoile, France) was performed to assess carbapenem hydrolysis.

### Epidemiological data and analysis

Epidemiological data of persons from whom *Enterobacter* spp. and *K. aerogenes* isolates were obtained from January 2012 until June 2019 were retrieved via the epidemiological questionnaire with demographic variables and predefined risk-factors that was part of the national CPE surveillance using a web-based system, called Type-Ned CPE. Starting from July 2019, it became mandatory to notify patients with CPE, and questionnaire data were collected by the Municipal Health Services and forwarded to RIVM through the national web-based notification system, called OSIRIS. When epidemiological data was available in both databases, Type-Ned data was used. Numbers and percentages for sex, sample material, reason of culture and risk details of the *Enterobacter* and *K. aerogenes* isolates per phenotype were calculated. The median and interquartile range (IQR) were calculated for age.

### Illumina next-generation sequencing

Illumina next-generation sequencing (NGS) was performed on all isolates for genotypic confirmation of the presence of Ambler class A to D β-lactamases. In brief, DNA was extracted using the DNeasy Blood & Tissue Kit (Qiagen, Hilden, Germany) until 2023. Afterwards, DNA was extracted using Maxwell RSC Cultured Cells DNA kit AS1620 (Promega, 17). DNA libraries were prepared using Illumina DNA Prep (Illumina, San Diego, CA, USA). Paired-end sequencing (2 x 150 bp) was performed on the Illumina NextSeq550 platform (Illumina, USA). Read quality analysis and *de novo* assembly was performed with Juno-assembly v2.0.2 pipeline (https://github.com/RIVM-bioinformatics/juno-assembly). NGS data was used for resistome and replicome analyses with the comprehensive antibiotic resistance database (CARD) (v6.0.3), AMRFinder (v3.11.26), ResFinder (v4.4.2) and PlasmidFinder (v2.1.6) software. A threshold of 95% was used for identity and 60% for the minimum length for ResFinder. Additionally, NGS data was used for classical multi-locus sequence typing (MLST), pan-genome MLST (pgMLST) (18), and whole genome multi-locus sequence typing (wgMLST) to detect genetic clustering. Genetic clusters for CIM+Carba-*Enterobacter* isolates were identified using pgMLST (18), and for CIM+Carba-*K. aerogenes* using wgMLST. The Enterobacter pgMLST scheme comprised 9,829 core genes derived from three reference genomes: 2,347 core genes from *E. roggenkampii* strain DSM 16690 (CP017184), 2,864 core genes from *E. hormaechei* subsp. *hoffmannii* strain DSM 14563 (CP017186) and 4,618 core genes from *E. cloacae* subsp. *cloacae* strain ATCC 13047 (CP001918). The *K. aerogenes* wgMLST scheme comprised 3,854 core and 911 accessory genes using the reference genome CP055904 and query genomes CP031756, CP041925, CP077265 and CP103664. Clusters were included if they contained at least two isolates originating from a minimum of two persons, and with a maximum allelic distance difference of 20 pgMLST/wgMLST alleles.

### Nanopore long-read sequencing

Long-read sequencing was performed on 99 isolates to determine the genomic locations (chromosome/plasmid) of genes encoding Ambler class C-type enzymes. High-molecular-weight DNA was isolated using an in-house developed protocol as described previously for isolates until 2023 (19, 20). From 2023 onwards, DNA was isolated using an automated nucleic acid extraction and purification platform from Maxwell (Promega, 17). The Maxwell RSC Cultured Cells DNA kit (AS1620), capable of isolating DNA up to 48 bacterial isolates per run. The manufacturer’s instructions were followed, except nuclease-free water was used instead of TE buffer to create the cell suspension, and RNase treatment was omitted. The Oxford Nanopore protocols SQK-LSK108, SQK-LSK109, SQK-RBK104 and SQK-RBK114 were used (Oxford Nanopore Technologies, https://community.nanoporetech.com). Until 2019, shearing was performed using g-TUBEs (Covaris) to obtain DNA fragment sizes of 8 kb for *Enterobacter/K. aerogenes* isolates. However, this shearing step was omitted for all isolates from 2019 onwards to obtain larger DNA fragments and simplify the protocol. For ligation, the SQK-LSK108 was used until September 2018, while SQK-LSK109 was used afterwards. Ligation protocols had DNA repaired using FFPE and end-repair kits (New England BioLabs), followed by ligation of barcodes with 1× bead clean up using AMPure XP (Beckman Coulter Nederland) as described in SQK-LSK108 and SQK-LSK109. Barcoded isolates were pooled, and sequencing adapters were added by ligation. The ligation protocols were followed until October 2020, after which only rapid protocols were used. In brief, barcoded transposome complexes were used to tagment the DNA, and simultaneous attaching a pair of barcodes. The final library was loaded onto a MinION flow cell, using FLO-MIN106 R9.4.1 for all isolates up to 2022, and FLO-MIN114 R10.4.1 for isolates from 2023 onwards. The length of a sequence run was 48 hours until 2020, after which it increased to 72 hours. Base calling and de-multiplexing were performed using Albacore for isolates until 2018, afterwards Guppy was used. Sequence data was extracted from the FAST5 files using Poretools until 2018 (21), while afterwards sequence data was extracted using NanoFilt [https://github.com/wdecoster/nanofilt] and Filtlong [https://github.com/rrwick/Filtlong]. Fifty base pairs were trimmed at both sides for ligation protocol isolates and eighty base pairs for rapid protocol isolates. Only reads larger than 5000-bp were used in further analyses. Illumina and Nanopore data were used in a hybrid assembly performed by Unicycler (22), which was operated using default settings and verbosity 2. The resulting contig files were annotated using Bakta 1.6.1 and were subsequently loaded into BioNumerics v8.1 for further analyses (23). Both raw NGS and Nanopore long-read sequence data are available at the sequence read archive (SRA; PRJEB35685, PRJNAB903550, PRJNA1076808 and PRJNA1122997).

### MASH-based *Enterobacter* spp. and *K. aerogenes* population analysis

Pairwise distances between the genome sequences, including available *Enterobacter* spp. and *K. aerogenes* reference genomes (NCBI Datasets Genome, Retrieved 29-01-2024), were calculated from NGS data using MASH v2.3 with a k-mer size k=21 and a sketch size s=5000 (24). MASH converts sequences into a MinHash sketch which was used to estimate the Jaccard index, p-value and MASH distance. The resulting network of genomes was visualized in Cytoscape v3.9.1 as Kruskal’s minimal spanning tree (25). Networks per (sub)species were created based on a MASH distance cut-off of 0.025 (Average Nucleotide Identity ∼97.5%) (Supplementary Figure S1) (24).

### Detection of ACT-28 protein in CIM supernatant

CIM supernatant was purified through centrifugation and concentrated using centrifugal filter units with a 10 kDa cutoff (Sigma-Aldrich, Missouri, United States). SDS sample buffer (Merck Millipore, Massachusetts, United States) was added to the supernatant, which was subsequently heated and analyzed by SDS-PAGE (stain-free, 4-20% gradient gels, 15 well [Bio-Rad, California, United States]) using 1x Tris/Glycine/SDS buffer (Bio-Rad, California, United States) and Precision Plus unstained protein standard (Bio-Rad). Nanoscale liquid chromatography coupled to tandem mass spectrometry (NanoLC-MS/MS) was performed by Alphalyse A/S (Odense, Denmark) on excised SDS-PAGE gel slices to determine protein identity. SignalP 5.0 (Technical University of Denmark DTU) was used to predict the length and cleavage site of the ACT-28 signal peptide. ProtParam (Expasy, Swiss Bioinformatics Resource Portal) was used to determine the molecular mass of ACT-28 in kDa.

### Ethics statement

The bacterial isolates belong to the MMLs participating in the Dutch national CPE surveillance program and were obtained as part of routine clinical care. Ethical approval and consent were not needed for the study, since it is based on surveillance data only. According to the Dutch Medical Research Involving Human Subjects Act (WMO) this study was exempt from review by an institutional review board.

## Results

### Carbapenemase-producing Enterobacterales from the Dutch national surveillance, 2012-2023

Between 2012-2023, the RIVM received 2,971 CPE isolates, and 49 non-CPE isolates were included in this study (n=3,020) (Table 1). The 2,971 isolates had a positive phenotypic confirmation for carbapenemase production by the CIM; termed CIM+, and/or a positive genotypic confirmation of the presence of major carbapenemase genes (IMP, KPC, NDM, OXA-48-like, and VIM); called Carba+. The majority of isolates were *K. pneumoniae* (*n*=1,202) and *E. coli* (*n=*993), followed by *E. cloacae* complex (*n=*351). A large proportion of the CIM+ *E. cloacae* complex and *K. aerogenes* (111/351; 31.6% and 13/25; 52.0%) isolates failed to identify a major carbapenemase gene by PCR; a phenotype/genotype termed CIM+Carba-(Table 1). This phenotype was found less frequently in *K. pneumoniae* (3/1,202; 0.3%) and *E. coli* (8/993; 0.8%), therefore the study is focused on *E. cloacae* complex and *K. aerogenes*. The dataset for further analyses included a total of 399 *E. cloacae* complex and *K. aerogenes* isolates that met the criteria of having available NGS data, consisting of 234 CIM+Carba+, 116 CIM+Carba- and 49 CIM-Carba-isolates (Table 1, Supplementary File 1). The CIM-Carba-group included 49 isolates of which 41 were *E. cloacae* complex and 8 were *K. aerogenes* isolates. The majority (95/116; 81.9%) of the CIM+Carba-*E. cloacae* complex and *K. aerogenes* isolates had MICs for meropenem that were below the clinical breakpoint of 2 mg/L for susceptibility according to EUCAST v15.0 (Table 2). The CIM+Carba+ and CIM-Carba-*E. cloacae* complex and *K. aerogenes* isolates were more often resistant to meropenem than the CIM+Carba-isolates (Table 2).

**Table 1.** Carbapenemase-producing Enterobacterales sent to RIVM for national surveillance between 2012 and 2023.

| Species | CIM+Carba+ | CIM+Carba- | CIM+Carba-<br>(%) | CIM-<br>Carba+ (*) | Total | CIM-Carba-<br>(non-CPE) |
| --- | --- | --- | --- | --- | --- | --- |
| *Klebsiella pneumoniae* | 1,194 | 3 | 0.2% | 5 | 1,202 | 0 |
| *Escherichia coli* | 982 | 8 | 0.8% | 3 | 993 | 0 |
| *Enterobacter cloacae* complex | 240 | 111 | 31.6% | 0 | 351 | 41 |
| *Citrobacter freundii* | 164 | 1 | 0.6% | 0 | 165 | 0 |
| *Proteus mirabilis* | 43 | 7 | 14.0% | 0 | 50 | 0 |
| *Klebsiella oxytoca* | 48 | 0 | 0.0% | 0 | 48 | 0 |
| *Klebsiella aerogenes* | 12 | 13 | 52.0% | 0 | 25 | 8 |
| *Serratia marcescens* | 19 | 5 | 20.8% | 0 | 24 | 0 |
| *Morganella morganii* | 15 | 3 | 16.7% | 0 | 18 | 0 |
| Other CPE | 93 | 2 | 2.1% | 0 | 95 | 0 |
| Total | 2,810 | 153 | 5.1% | 8 | 2,971 | 49 |

\*Inactive carbapenemases due to point mutations, premature stopcodons or frame shifts.

**Table 2.**
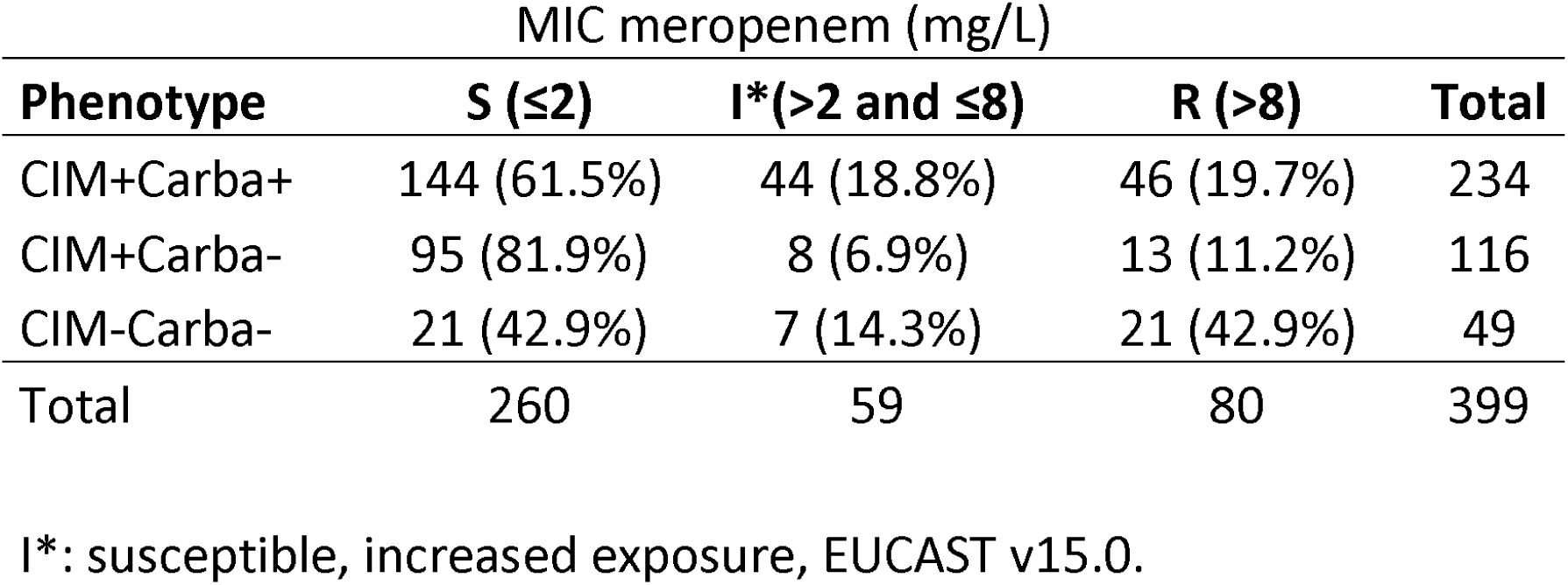
Minimum inhibitory concentration for meropenem of 399 *E. cloacae* complex/*K. aerogenes* from different phenotypes.

### Epidemiological characteristics of persons carrying and/or infected with *E. cloacae* complex/*K. aerogenes*

The 399 *E. cloacae* complex and *K. aerogenes* isolates belong to 399 persons, of whom 217 were male (54.4%), 179 were female (44.9%) and for three persons it was unknown (0.8%) (Table 3). The median age was 67 years old. CIM+Carba+ and CIM+Carba-*E. cloacae* complex/*K. aerogenes* isolates were cultured mostly from swabs (69.7%, 35.3.%, respectively), followed by urine (9.4%, 34.5%, respectively). Of the persons with a CIM+Carba+ isolate, 62.4% were sampled as part of routine or targeted screening because of increased risk of CPE carriage, while for persons with a CIM+Carba-isolate this was 22.4%. Persons with a CIM+Carba+ *E. cloacae* complex/*K. aerogenes* isolate were sampled for diagnostic purposes in 16.2% of the cases, while this was 40.5% for persons with a CIM+Carba-isolate. Of the individuals from whom CIM+Carba+ and CIM+Carba-*E. cloacae* complex/*K. aerogenes* isolates were obtained, 36.3% and 50.0% had no known risk factors, respectively. Of the persons with a CIM+Carba+ *E. cloacae* complex/*K. aerogenes* isolate, 30.3% were admitted to a foreign hospital for more than 24 hours, less than 2 months before sampling, indicating that a large proportion of the person were probably colonized abroad. In contrast, for persons with a CIM+Carba-sample this was only 6.9%, suggesting these were primarily contracted in the Netherlands.

**Table 3.** Characteristics of persons with *E. cloacae* complex/*K. aerogenes*, divided per phenotype/genotype.

|  | All |  | CIM+Carba+ |  | CIM+Carba- |  | CIM-Carba- |  |
| --- | --- | --- | --- | --- | --- | --- | --- | --- |
| Isolates | 399 |  | 234 |  | 116 |  | 49 |  |
| Sending institutions | 47 |  | 42 |  | 37 |  | 27 |  |
| *Sex* |  |  |  |  |  |  |  |  |
| Male | 217 | 54.4% | 137 | 58.5% | 58 | 50.0% | 22 | 44.9% |
| Female | 179 | 44.9% | 96 | 41.0% | 58 | 50.0% | 25 | 51.0% |
| Unknown | 3 | 0.8% | 1 | 0.4% | 0 | 0% | 2 | 4.1% |
| *Age* |  |  |  |  |  |  |  |  |
| Median age (IQ1-IQ3) | 67 | (52-78) | 67 | (52-78) | 66 | (52-76) | 68 | (53.5-78) |
| *Sample material* |  |  |  |  |  |  |  |  |
| Blood | 13 | 3.3% | 5 | 2.1% | 7 | 6.0% | 1 | 2.0% |
| Pus/puncture/biopsy | 22 | 5.5% | 8 | 3.4% | 10 | 8.6% | 2 | 4.1% |
| Sputum/BAL | 20 | 5.0% | 9 | 3.8% | 7 | 6.0% | 4 | 8.2% |
| Swab of throat, nose, perineum, and/or rectum | 222 | 55.6% | 163 | 69.7% | 41 | 35.3% | 18 | 36.7% |
| Urine | 73 | 18.3% | 22 | 9.4% | 40 | 34.5% | 11 | 22.4% |
| Wound/ulcer/superficial infection | 23 | 5.8% | 14 | 6.0% | 6 | 5.2% | 3 | 6.1% |
| Other human material | 8 | 2.0% | 5 | 2.1% | 1 | 0.9% | 4 | 8.2% |
| Unknown | 18 | 4.5% | 8 | 3.4% | 4 | 3.4% | 6 | 12.2% |
| *Reason for sampling* |  |  |  |  |  |  |  |  |
| Screening | 173 | 43.4% | 146 | 62.4% | 26 | 22.4% | 1 | 2.0% |
| Diagnostic | 87 | 21.8% | 38 | 16.2% | 47 | 40.5% | 2 | 4.1% |
| Unknown | 139 | 34.8% | 50 | 21.4% | 43 | 37.1% | 46 | 93.9% |
| *Identified risk factors* |  |  |  |  |  |  |  |  |
| Admitted to a foreign hospital for >24 hours, <2 months ago | 79 | 19.8% | 71 | 30.3% | 8 | 6.9% | 0 | 0.0% |
| Contact with a foreign hospital >2 months <1 year ago | 12 | 3.0% | 11 | 4.7% | 1 | 0.9% | 0 | 0.0% |
| Known carrier of CPE | 8 | 2.0% | 6 | 2.6% | 2 | 1.7% | 0 | 0.0% |
| Known CPE outbreak in care facility | 4 | 1.0% | 4 | 1.7% | 0 | 0.0% | 0 | 0.0% |
| Previous admission to other Dutch institution with ongoing CPE outbreak | 7 | 1.8% | 6 | 2.6% | 1 | 0.9% | 0 | 0.0% |
| Travel to a foreign country without hospital visit or admission in the past 12 months | 5 | 1.3% | 3 | 1.3% | 2 | 1.7% | 0 | 0.0% |
| No known risk factors | 146 | 36.6% | 85 | 36.3% | 58 | 50.0% | 3 | 6.1% |
| Unknown | 138 | 34.6% | 48 | 20.5% | 44 | 37.9% | 46 | 93.9% |

### MASH revealed 18 genogroups comprising *Enterobacter* spp./*K. aerogenes* in the Netherlands

MASH analysis on NGS data of 399 genomes in combination with RefSeq genomes of curated *Enterobacter* spp. and *K. aerogenes* enabled species determination, and revealed 18 genomic groups (GGs). These groups include 15 GGs corresponding to confirmed *Enterobacter* species and three GGs associated with *K. aerogenes* (Fig. 1, Table 4, Supplementary Figure S1). Generally, the isolates per-genogroup-per-phenotype were restricted to specific STs (Table 5). The largest GG comprised *E. hormaechei* subsp. *steigerwaltii* (GG01e, *n=*168) and is associated with multilocus sequence typing (MLST) sequence types (ST) ST45, ST114, ST121, ST171 and ST182. The group was followed by GG02e which consisted of *E. kobei* (*n=*63, ST32, ST56, ST122, ST125 and ST591) and *E. hormaechei* (GG03e, *n=*53, ST78), *E. roggenkampii* (GG04e, *n=*30, ST165, ST1059) and *K. aerogenes* (GG01k, *n=*26, ST93). Seven smaller GGs comprised: *E. cloacae* (GG05e, *n=*20, ST135), *E. asburiae* (GG06e, *n=*7, ST456), *E. ludwigii* (GG07e, *n=*6), *E. bugandensis* (GG08e, *n=*3), *E. mori* (GG09e, *n=*1), *E. quasiroggenkampii* (GG10e, *n=*1) and *E. sichuanensis* (GG11e, *n=*1), respectively. Eighteen isolates, from 6 genomic groups (GG12e-GG15e, GG02k-GG03k), and 2 single isolates were not associated with a RefSeq genome. Fourteen RefSeq genomes did not associate with any Dutch surveillance *Enterobacter/K. aerogenes* isolate. The CIM+Carba+, CIM+Carba- and CIM-Carba-phenotypes were non-randomly distributed among *Enterobacter* spp. The three largest CIM+Carba+ groups were comprised of *E. hormaechei* subsp. *steigerwaltii*, *E. hormaechei* and *E. cloacae* (Fig. 1). The three relatively largest CIM+Carba-groups were *E. kobei, E. roggenkampii* and *E. ludwigii* and will therefore be the focus for further in-depth analyses. Based on pgMLST, five small (≤4 isolates/cluster) genetic clusters (≥2 isolates varying ≤20 pgMLST alleles) comprised 13 CIM+Carba-*Enterobacter* isolates from 13 patients, while seventeen clusters containing 109 CIM+Carba+ *Enterobacter* spp. isolates with major carbapenemase genes. Four CIM+Carba-clusters comprised eleven *E. kobei* isolates, while the fifth cluster consisted of two *E. roggenkampii* isolates. Two of the 5 CIM+Carba-clusters represented unconfirmed nosocomial spread without epidemiological link, while 3/5 CIM+Carba-clusters were from isolates obtained over multiple years (Supplementary Figure S2). wgMLST analysis of CIM+Carba-*K. aerogenes* revealed no genetic clusters, suggesting no nosocomial spread (Supplementary Figure S3).

**Figure 1.**
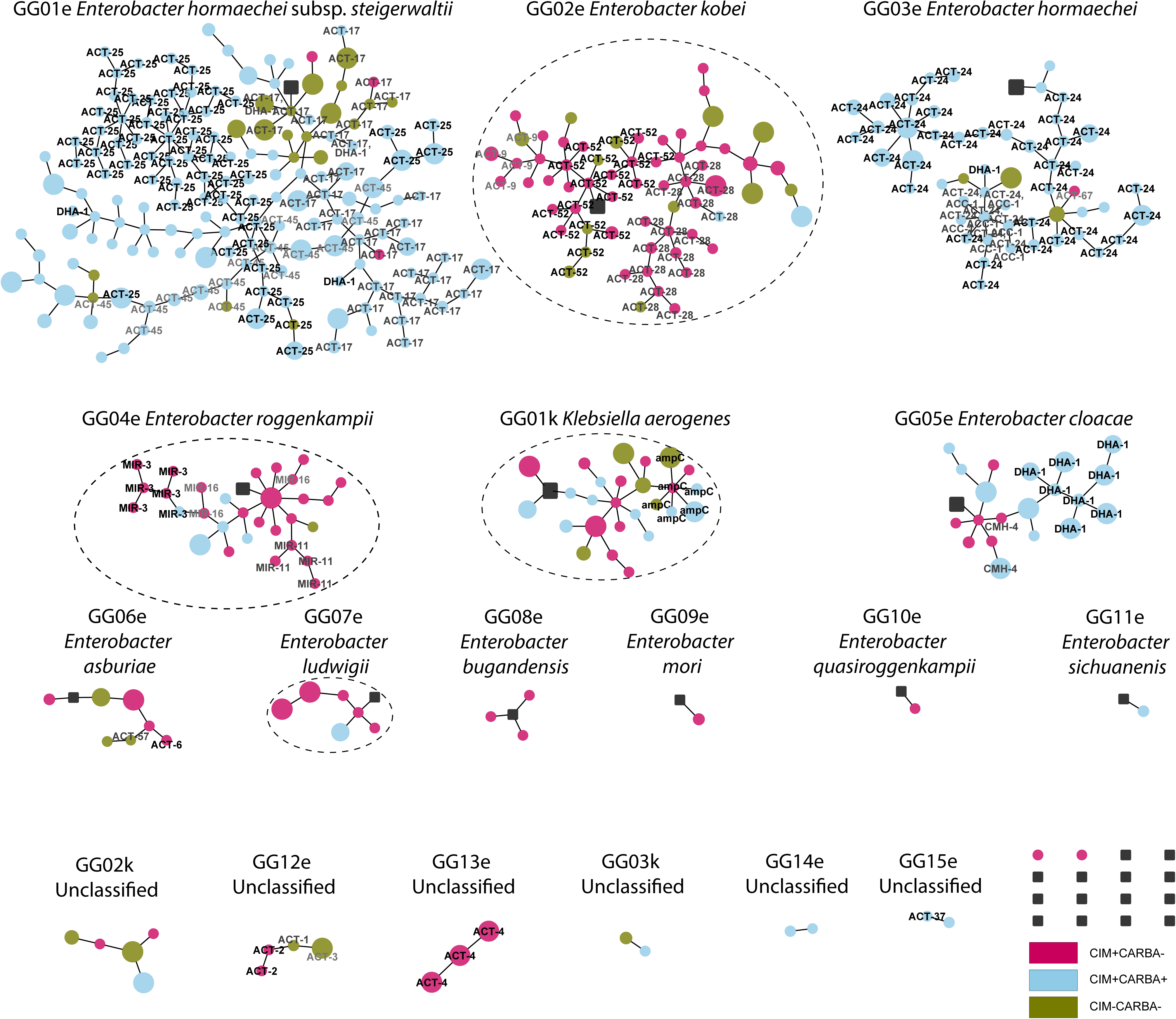
*Enterobacter* spp. and *K. aerogenes* population in the Netherlands associates with CIM phenotype, Ambler class C β-lactamases and resistance to meropenem. MASH analyses of 366 *Enterobacter* spp. and 25 *K. aerogenes* isolates revealing fifteen *Enterobacter* and three *K. aerogenes* genogroups. Each dot represents an isolate. The color of the dot shows different phenotypes: blue (CIM+Carba+), magenta (CIM+Carba-) and green (CIM-Carba-). The top 3 Ambler class C β-lactamase genes per species are shown. The size of dots show susceptibility to meropenem; how larger the dot, the higher the MIC of meropenem. Each black square represents a reference genome isolate.

**Table 4.** MASH revealing fifteen *Enterobacter* spp. and three *K. aerogenes* genomic groups belonging to different phenotypes.

| Genomic Group (GG) | CIM+Carba+ | CIM+Carba- | CIM-Carba- | Total |
| --- | --- | --- | --- | --- |
| GG01e *Enterobacter hormaechei* subsp. *steigerwaltii* | 146 | 3 | 19 | 168 |
| GG02e *Enterobacter kobei* | 2 | 48 | 13 | 63 |
| GG03e *Enterobacter hormaechei* | 49 | 1 | 3 | 53 |
| GG04e *Enterobacter roggenkampii* | 5 | 24 | 1 | 30 |
| GG05e *Enterobacter cloacae* | 14 | 6 | 0 | 20 |
| GG06e *Enterobacter asburiae* | 0 | 4 | 3 | 7 |
| GG07e *Enterobacter ludwigii* | 1 | 5 | 0 | 6 |
| GG08e *Enterobacter bugandensis* | 0 | 3 | 0 | 3 |
| GG09e *Enterobacter mori* | 0 | 1 | 0 | 1 |
| GG10e *Enterobacter quasiroggenkampii* | 0 | 1 | 0 | 1 |
| GG11e *Enterobacter sichuanensis* | 1 | 0 | 0 | 1 |
| GG12e Unclassified | 0 | 2 | 2 | 4 |
| GG13e Unclassified | 0 | 3 | 0 | 3 |
| GG14e Unclassified | 2 | 0 | 0 | 2 |
| GG15e Unclassified | 2 | 0 | 0 | 2 |
| GG01k *Klebsiella aerogenes* | 10 | 11 | 5 | 26 |
| GG02k Unclassified | 1 | 2 | 2 | 5 |
| GG03k Unclassified | 1 | 0 | 1 | 2 |
| Other | 0 | 2 | 0 | 2 |
| Total | 234 | 116 | 49 | 399 |

**Table 5.** Association of most common MLST sequence types with the *Enterobacter* spp. and *K. aerogenes* genomic groups in the Netherlands.

| *Enterobacter* spp. | Genomic Group (GG) | MLST ST | CIM+Carba+ | CIM+Carba- | CIM-Carba- | Total |
| --- | --- | --- | --- | --- | --- | --- |
| *E. hormaechei* subsp. *steigerwaltii* | GG01e | 45 | 8 | 0 | 0 | 8 |
|  |  | 114 | 16 | 0 | 0 | 16 |
|  |  | 121 | 50 | 0 | 1 | 51 |
|  |  | 171 | 8 | 0 | 0 | 8 |
|  |  | 182 | 6 | 0 | 0 | 6 |
|  |  | Other | 58 | 3 | 18 | 79 |
| *E. kobei* | GG02e | 32 | 0 | 10 | 4 | 14 |
|  |  | 56 | 0 | 3 | 0 | 3 |
|  |  | 122 | 0 | 4 | 2 | 6 |
|  |  | 125 | 1 | 14 | 2 | 17 |
|  |  | 591 | 0 | 2 | 1 | 3 |
|  |  | Other | 1 | 15 | 4 | 20 |
| *E. hormaechei* | GG03e | 78 | 41 | 0 | 1 | 42 |
|  |  | Other | 8 | 1 | 2 | 11 |
| *E. roggenkampii* | GG04e | 165 | 0 | 4 | 0 | 4 |
|  |  | 1059 | 0 | 3 | 0 | 3 |
|  |  | Other | 5 | 17 | 1 | 23 |
| *E. cloacae* | GG05e | 456 | 10 | 0 | 0 | 10 |
|  |  | Other | 4 | 6 | 0 | 10 |
| *E. asburiae* | GG06e | Other | 0 | 4 | 3 | 7 |
| *E. ludwigii* | GG07e | Other | 1 | 5 | 0 | 6 |
| *E. bugandensis* | GG08e | Other | 0 | 3 | 0 | 3 |
| *E. mori* | GG09e | Other | 0 | 1 | 0 | 1 |
| *E. quasiroggenkampii* | GG10e | Other | 0 | 1 | 0 | 1 |
| *E. sichuanensis* | GG11e | Other | 1 | 0 | 0 | 1 |
| Unclassified | GG12e | Other | 0 | 2 | 2 | 4 |
| Unclassified | GG13e | Other | 0 | 3 | 0 | 3 |
| Unclassified | GG14e | Other | 2 | 0 | 0 | 2 |
| Unclassified | GG15e | Other | 2 | 0 | 0 | 2 |
| *K. aerogenes* | GG01k | 93 | 3 | 1 | 2 | 6 |
|  |  | 135 | 0 | 3 | 0 | 3 |
|  |  | Other | 7 | 7 | 3 | 17 |
| Unclassified | GG02k | Other | 1 | 2 | 2 | 5 |
| Unclassified | GG03k | Other | 1 | 0 | 1 | 2 |
| Other | non-GG | Other | 0 | 2 | 0 | 2 |
| Total |  |  | 234 | 116 | 49 | 399 |

### Resistome and replicome diversity among *Enterobacter* spp. and *K. aerogenes*

The resistome and replicome were enriched in Ambler class A-D β-lactamase genes and plasmid replicons among CIM+Carba+ *Enterobacter* spp. and *K. aerogenes*, while CIM+Carba- and CIM-Carba-isolates were mostly devoid of β-lactamase genes and replicons (Fig. 2 and Fig. 3). Among CIM+Carba+ isolates, Ambler class C enzymes were found in 99.1% of the isolates, followed by class A (81.2%), D (81.2%) and B (37.6%) enzymes, respectively. Major β-lactamase gene *bla*_OXA-48_ was found most (54.7%) among CIM+Carba+ isolates, followed by *bla*_OXA-1_ (50.9%), *bla*_TEM-1_ (50.0%), and *bla*_CTX-M-15_ (46.6%), respectively. The majority of CIM+Carba+ *E. hormaechei* subsp. *steigerwaltii* harbored *bla*_TEM-1_ (70.5%), *bla*_CTX-M-15_ (64.4%), *bla*_OXA-1_ (63.7%) and the *bla*_OXA-48_ gene (61.6%), respectively. *bla*_ACT-24_ and *bla*_OXA-48_ were found in a large proportion (85.7% and 63.3%, respectively) of CIM+Carba+ *E. hormaechei*, while CIM+Carba+ *E. cloacae* harbored mostly *bla*_KPC-2_ (78.6%) and *bla*_OXA-1_ (71.4%), respectively. All CIM+Carba+ isolates harbored at least one plasmid replicon, and 95.3% of the isolates had a 100%-identity match with a plasmid replicon. The largest proportion (98.3%) of CIM+Carba-isolates harbored Ambler class C-type enzymes, followed by Class A (15.5%) and D (1.7%), respectively. The three relatively largest CIM+Carba-groups in the Netherlands were *E. kobei, E. roggenkampii* and *E. ludwigii*. CIM+Carba-*E. kobei* harbored only *bla*_ACT_ alleles, with *bla*_ACT-28_ and *bla*_ACT-52_ found most often in 31.3% and 27.1% of the cases, respectively. *E. roggenkampii* harbored only *bla*_MIR_-type alleles, of which *bla*_MIR-3_ and *bla*_MIR-11_ were found most often (20.8% and 16.7%, respectively). *E. ludwigii* harbored various *bla*_ACT_-like alleles. 60.3% of the CIM+Carba-isolates harbored a plasmid replicon, while only 27.6% of the CIM+Carba-isolates had a 100%-identity match with a plasmid replicon. Ninety-eight percent of the CIM-Carba-isolates harbored Ambler class C enzymes, followed by Ambler class A (51.0%) and D (4.1%), respectively. 71.4% of the CIM-Carba-isolates harbored a plasmid replicon, and 55.1% of the CIM-Carba-isolates had a 100%-identity match with a plasmid replicon. The CIM+Carba- and CIM-Carba-isolates with Class A and D enzymes had no known major IMP, KPC, NDM, OXA-48-like, and VIM carbapenemase enzymes.

**Figure 2.**
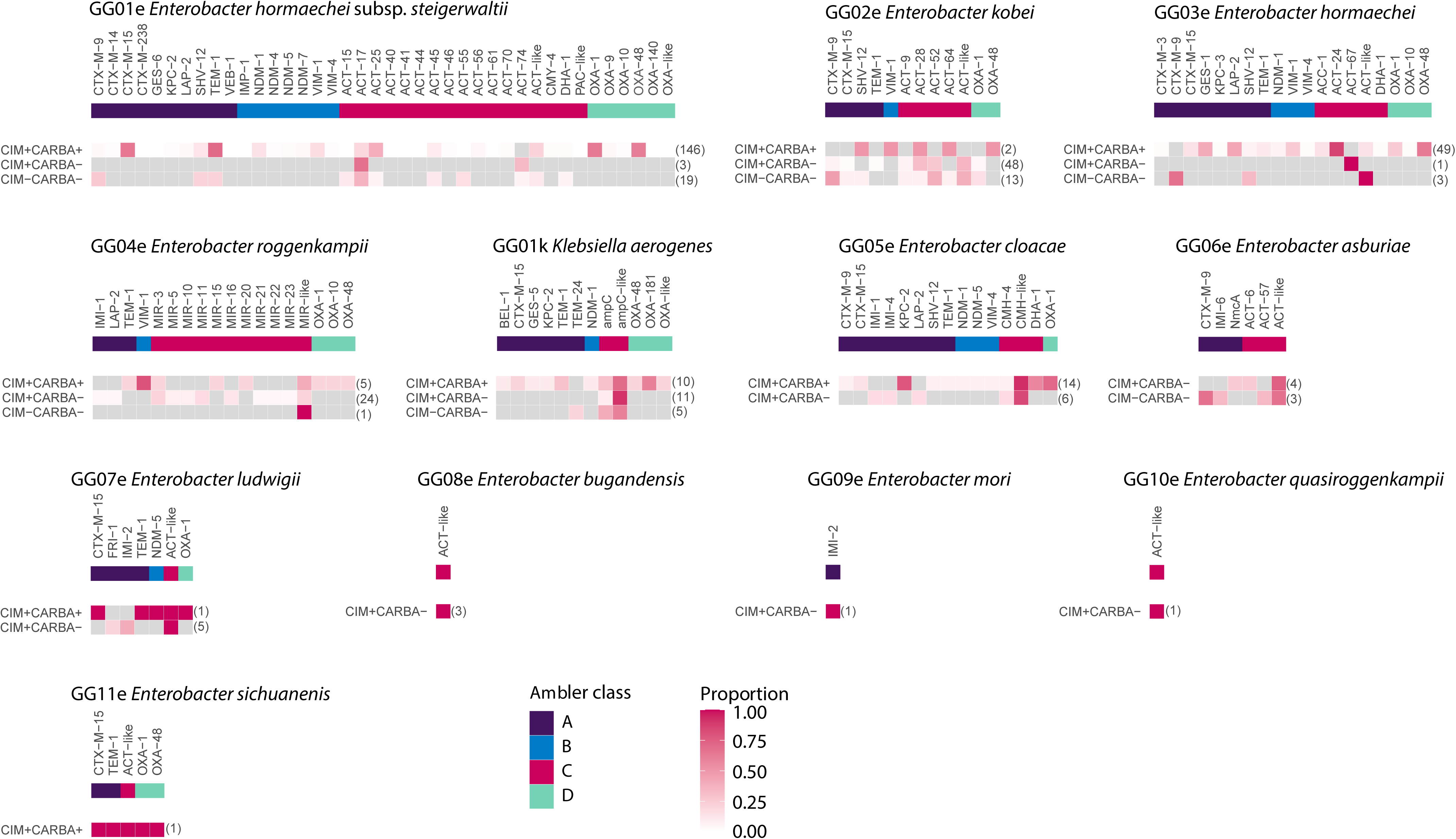
Resistome diversity among *Enterobacter* spp. and *K. aerogenes* population. The resistome of eleven *Enterobacter* species and *K. aerogenes* are shown per genogroup, and per phenotype. The upper bar reveals the presence of different β-lactamase genes among various Ambler classes. Purple represent the Ambler class A, blue encompasses class B, pink belongs to class C and green entails the class D enzymes. The intensity of a box is calculated by number of present beta-lactamase genes divided by number of isolates per species per phenotype - found between brackets. A grey box indicates that no allele was found.

**Figure 3.**
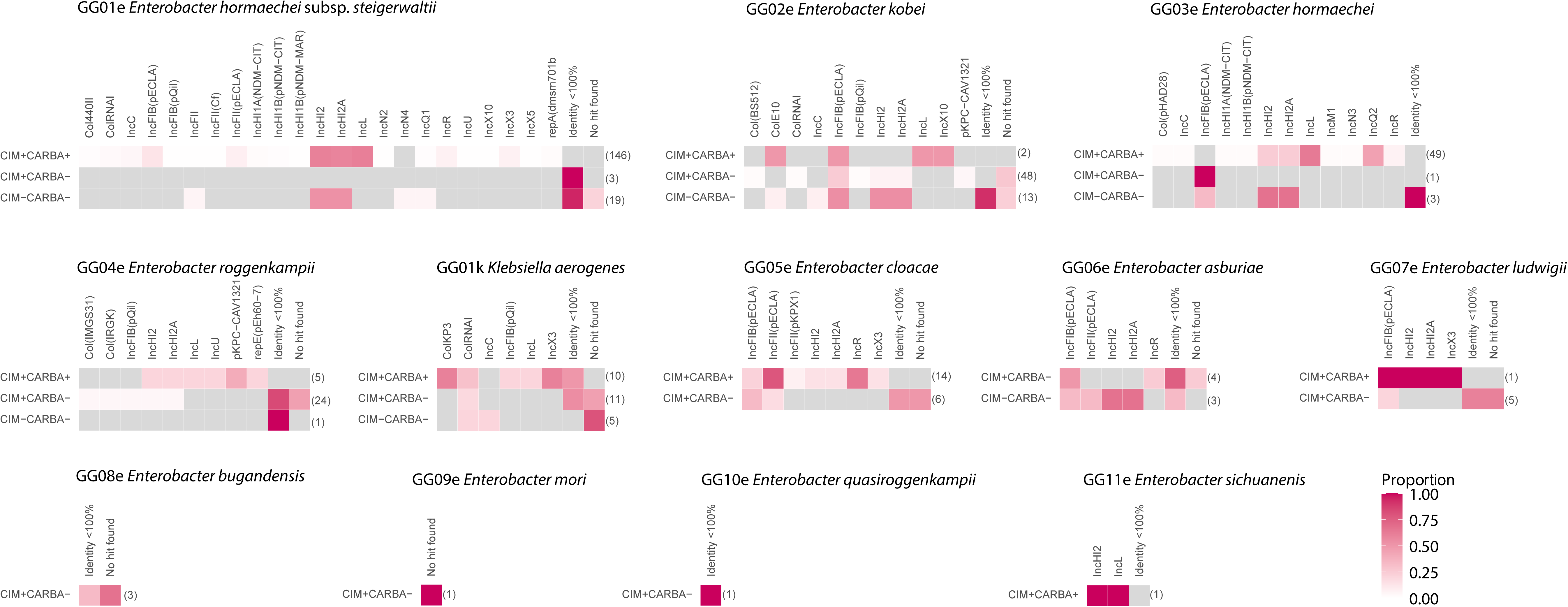
Replicome diversity among *Enterobacter* spp. and *K. aerogenes* population. The replicome of eleven *Enterobacter* species and *K. aerogenes* are shown per genogroup, and per phenotype. The upper bar reveals the screened plasmids and integrons. The intensity of a box is calculated by number of present replicomes divided by number of isolates per species per phenotype - found between brackets. A grey box indicates that no replicon was found.

### Distribution of Class C β-lactamase genes among *Enterobacter* spp./*K. aerogenes* in the Netherlands

CARD analysis for *Enterobacter* isolates and AMRFinder analysis for *K. aerogenes* of the NGS-derived resistomes revealed that 394 out of 399 isolates harbored one or more Ambler class C β-lactamase genes (*n*=418), these included genes such as *bla*_ACT_, *bla*_CMH_, *bla*_CMY_, *bla*_DHA_ and *bla*_MIR_ (Fig. 1, Table 6). Specific variant class C *bla*_ACT_-type alleles were restricted to specific genomic groups (Fig. 1 and Fig. 2). *bla*_ACT-9_-, *bla*_ACT-28_-, *bla*_ACT-52_-, *bla*_MIR-3_- and *bla*_MIR-11_-alleles (100% identity) were enriched (>80.0%) in the CIM+Carba-group comprising *E. kobei*, *E. roggenkampii* and *E. ludwigii* compared to the CIM+Carba+ group (Table 6). *bla*_ACT-17,_ *bla*_ACT-25_, *bla*_ACT-45_, *bla*_ACT-56_, and *bla*_DHA-1_ alleles were associated with CIM+Carba+ *E. hormaechei* subsp. *steigerwaltii* isolates from GG01e, while *bla*_ACT-9_, *bla*_ACT-28_ and *bla*_ACT-52_ alleles were associated with CIM+Carba-*E. kobei* from GG02e. *bla*_ACC-1_ and *bla*_ACT-24_ were associated with CIM+Carba+ *E. hormaechei* isolates from GG03e. *bla*_MIR-3_ and *bla*_MIR-11_ alleles were associated with CIM+Carba-*E. roggenkampii* from GG04e, while the *bla*_DHA-1_ allele was associated with CIM+Carba+ *E. cloacae* from GG05e, like GG01e. The majority (79.6%) of CIM-Carba-Ambler class C genes were *bla*_ACT_-like alleles, with *bla*_ACT-17_ found most frequently. The five isolates which did not harbor Ambler class C β-lactamase genes were one isolate from GG03e *E. hormaechei,* one *E. mori* isolate from GG09e, two isolates from GG14e were unclassified, and one single unclassified isolate. The other GGs were associated with other *bla*_ACT_-like, *bla*_CMH_-like and *bla*_DHA_-like alleles. The CarbaNP test was performed on *E. kobei, E. roggenkampii* and *E. ludwigii* isolates, along with the CIM, revealing a positive CarbaNP in 32/63 (50.8%), 5/6 (83.3%), and 25/30 (83.3%) of the isolates, respectively (Fig. 4). In comparison to the CIM, the CarbaNP test is less frequently positive. However, no isolate which was CIM negative, yielded a positive CarbaNP. Analysis of the CIM supernatant of an exemplaric *E. kobei* GG02e isolate containing *bla*_ACT-28_ revealed the presence of a dominant protein on reducing SDS-PAGE, which was absent in an *Enterobacter* isolate lacking *bla*_ACT-28_. Excision of the protein product and subsequent nano-LC-MS/MS protein identity determination demonstrated the presence of polypeptides corresponding to a processed and overproduced ACT-28 enzyme (Supplementary Figure S4).

**Figure 4.**
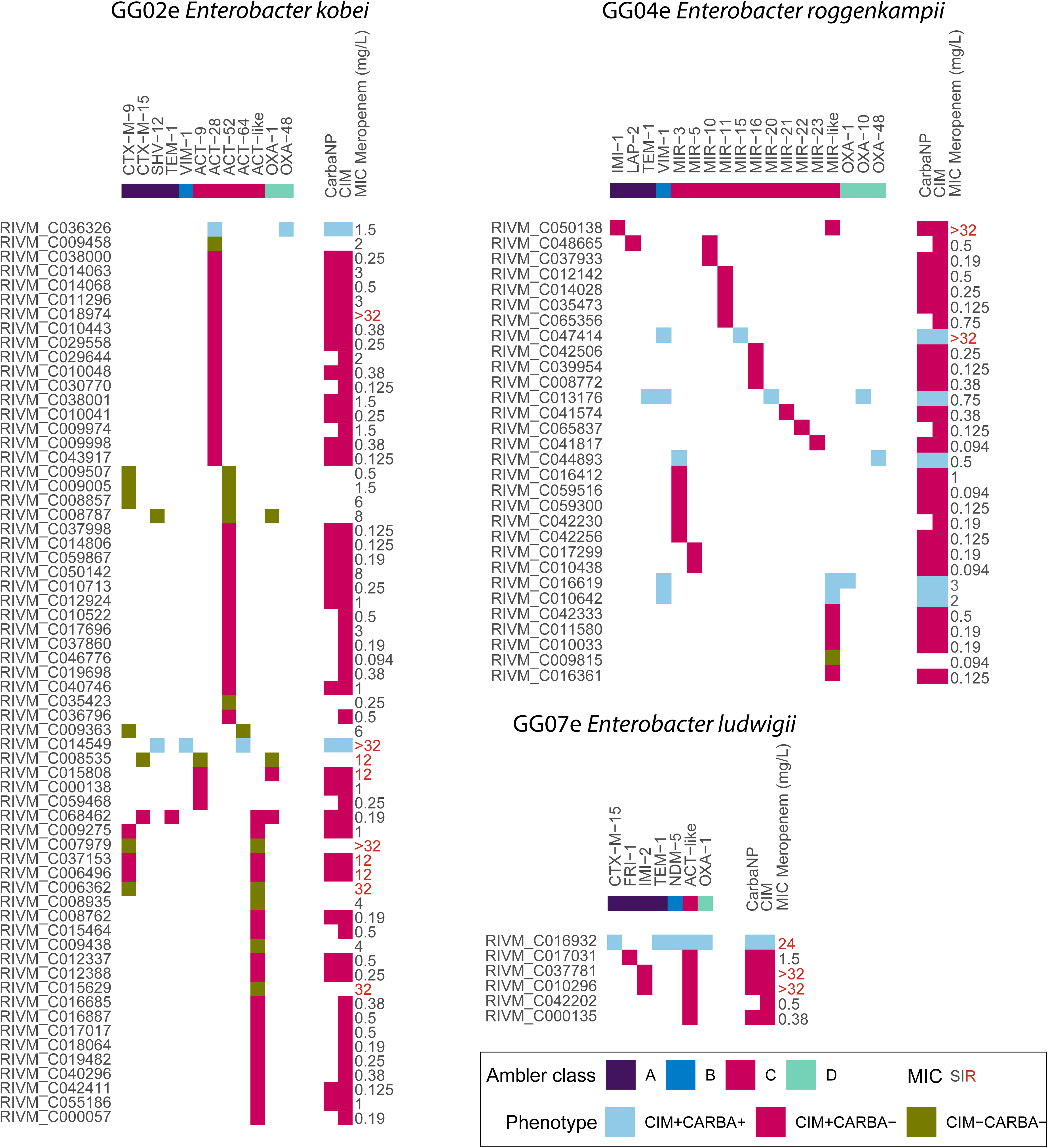
Phenotype/genotype plot of CIM+Carba-*E. kobei*, *E. roggenkampii* and *E. ludwigii*. The phenotype and genotype of 68 *E. kobei*, 30 *E. roggenkampii* and six *E. ludwigii* isolates are shown. For each isolate the presence of β-lactamase genes are indicated and the CarbaNP and/or CIM result when carbapenem hydrolysis and/or carbapenemase production phenotype occurred. MIC for meropenem is given in mg/L.

**Table 6.**
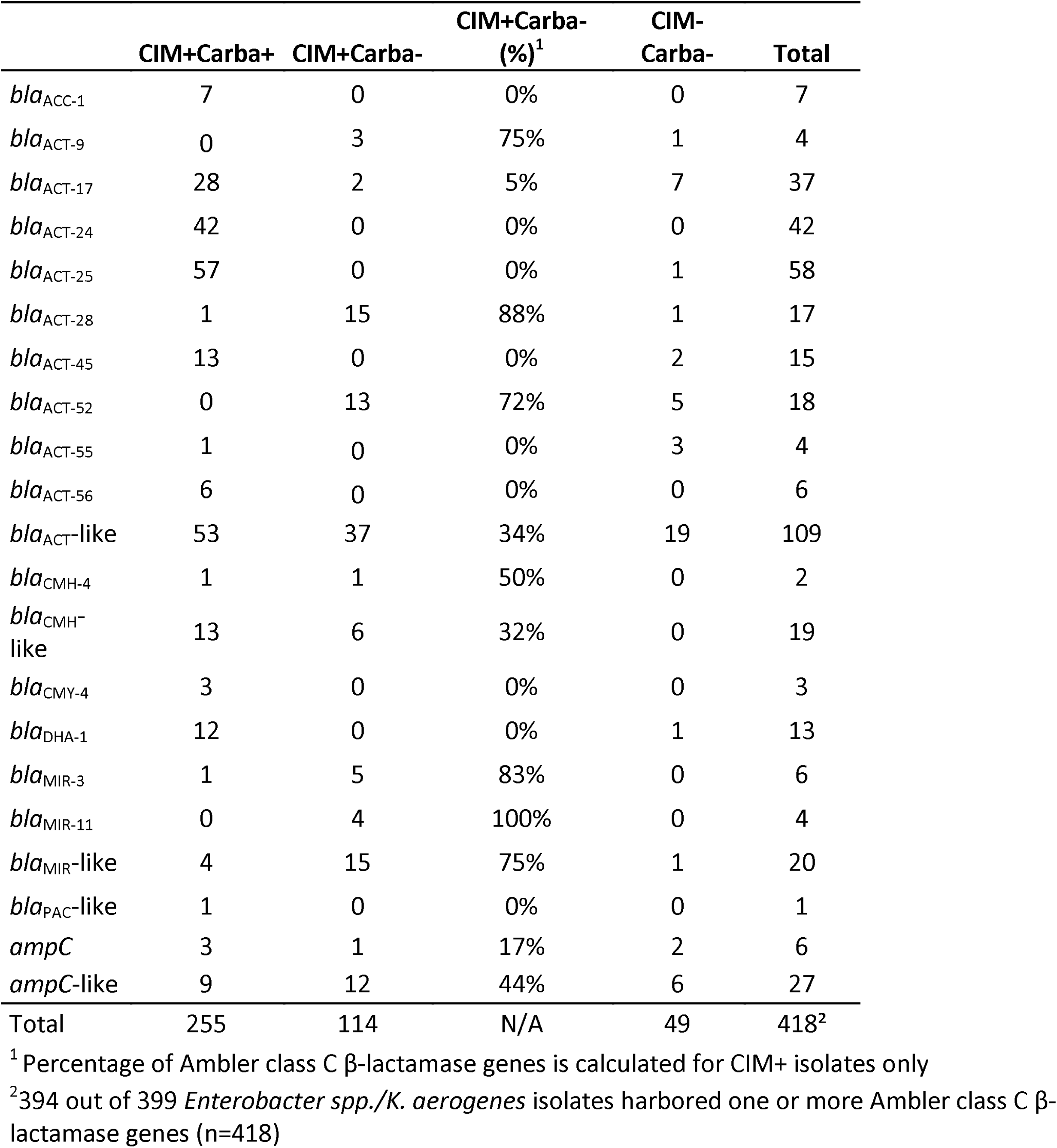
Distribution of Ambler class C β-lactamase genes in *Enterobacter* spp./*K. aerogenes*.

|  | CIM+Carba+ | CIM+Carba- | CIM+Carba-<br>(%)<sup>1</sup> | CIM-<br>Carba- | Total |
| --- | --- | --- | --- | --- | --- |
| *bla*<sub>ACC-1</sub> | 7 | 0 | 0% | 0 | 7 |
| *bla*<sub>ACT-9</sub> | 0 | 3 | 75% | 1 | 4 |
| *bla*<sub>ACT-17</sub> | 28 | 2 | 5% | 7 | 37 |
| *bla*<sub>ACT-24</sub> | 42 | 0 | 0% | 0 | 42 |
| *bla*<sub>ACT-25</sub> | 57 | 0 | 0% | 1 | 58 |
| *bla*<sub>ACT-28</sub> | 1 | 15 | 88% | 1 | 17 |
| *bla*<sub>ACT-45</sub> | 13 | 0 | 0% | 2 | 15 |
| *bla*<sub>ACT-52</sub> | 0 | 13 | 72% | 5 | 18 |
| *bla*<sub>ACT-55</sub> | 1 | 0 | 0% | 3 | 4 |
| *bla*<sub>ACT-56</sub> | 6 | 0 | 0% | 0 | 6 |
| *bla*<sub>ACT-like</sub> | 53 | 37 | 34% | 19 | 109 |
| *bla*<sub>CMH-4</sub> | 1 | 1 | 50% | 0 | 2 |
| *bla*<sub>CMH-like</sub> | 13 | 6 | 32% | 0 | 19 |
| *bla*<sub>CMY-4</sub> | 3 | 0 | 0% | 0 | 3 |
| *bla*<sub>DHA-1</sub> | 12 | 0 | 0% | 1 | 13 |
| *bla*<sub>MIR-3</sub> | 1 | 5 | 83% | 0 | 6 |
| *bla*<sub>MIR-11</sub> | 0 | 4 | 100% | 0 | 4 |
| *bla*<sub>MIR-like</sub> | 4 | 15 | 75% | 1 | 20 |
| *bla*<sub>PAC-like</sub> | 1 | 0 | 0% | 0 | 1 |
| *ampC* | 3 | 1 | 17% | 2 | 6 |
| *ampC-like* | 9 | 12 | 44% | 6 | 27 |
| Total | 255 | 114 | N/A | 49 | 418<sup>2</sup> |

<sup>1</sup> Percentage of Ambler class C β-lactamase genes is calculated for CIM+ isolates only
<sup>2</sup>394 out of 399 *Enterobacter* spp./*K. aerogenes* isolates harbored one or more Ambler class C β-lactamase genes (n=418)

### Genomic location of Ambler class C-type genes in *E. kobei and E. roggenkampii*

Ninety-nine isolates belonged to *E. kobei*, *E. roggenkampii* and *E. ludwigii* and were sequenced with both NGS and Nanopore long-read sequencing. Among the *E. kobei* and *E. roggenkampii* isolates, nineteen circular hybrid assemblies were obtained, while linear assemblies were omitted in this study (Supplementary Table S1). In all nineteen hybrid assemblies, Ambler class C β-lactamase genes, including *bla*_ACT-9_, *bla*_ACT-28_, *bla*_ACT-51_, *bla*_ACT-52_, *bla*_MIR-3_, *bla*_MIR-10_ *bla*_MIR-16_ and *bla*_MIR-22_ were located on the chromosome. The chromosome length of *E. kobei* varied between 4,708.241 and 5,042.444 bp, and the %GC content was between 54.8% and 55.1%. The chromosome length of *E. roggenkampii* varied between 4,597.662 to 4,854.898 bp, and the %GC content was between 55.9% and 56.1%. No circular hybrid assemblies were obtained for *E. ludwigii*.

## Discussion

The *Enterobacter* spp. population submitted to the CPE surveillance in the Netherlands between 2012-2023 is very diverse. *E. hormaechei* subsp. *steigerwaltii* occurs most often and consisted predominantly of carbapenemase-producing isolates with known carbapenemase genes, which is in accordance with a previous study (8). The largest group *E. hormaechei* subsp. *steigerwaltii* is followed by *E. kobei, E. hormaechei* and *E. roggenkampii.* Almost all (98.7%) of the *Enterobacter* isolates submitted to the Dutch national CPE surveillance carry Ambler class C-type β-lactamases, in line with an earlier study (11, 26). However, a large proportion of the *Enterobacter* isolates (31.6%) and *K. aerogenes* (52.0%) obtained in the Dutch national CPE surveillance was positive in the CIM phenotypical test for carbapenemase production in the absence of major carbapenemase genes (CIM+Carba-). *E. kobei, E. roggenkampii*, *E. ludwigii*, and *K. aerogenes* are species with this dominant CIM+Carba-phenotype. Our study showed that various class C-type β-lactamase genes, including *bla*_ACT-28_, *bla*_ACT-52_ and *bla*_MIR-3_, *bla*_MIR-11_ or *ampC*, were associated with the CIM+Carba-phenotype observed in *E. kobei, E. roggenkampii, E. ludwigii*, and *K. aerogenes* isolates, in line with a recent study (26). These isolates showed for a majority of CIM+ isolates also a positive CarbaNP test, and thus proof of hydrolysis. 81.9% of CIM+Carba-isolates had a meropenem susceptibility of <2 mg/L *in vitro*. The CIM+Carba-isolates were more frequently identified in samples taken for diagnostic purposes than those taken for routine or targeted screening in high risk groups. Some CIM+Carba-isolates that exhibited high MICs for meropenem, are likely due to the presence of *bla*_IMI_. In contrast, CIM+Carba+ isolates were found mostly in samples obtained for screening purposes, with 61.5% having a meropenem susceptibility of <2 mg/L. This may suggest that a large portion of the CIM+Carba+ *Enterobacter* spp./*K. aerogenes* isolates were acquired abroad, while CIM+Carba-isolates circulate endemically within the Netherlands. Given the higher prevalence levels of CPE in many other countries, the increased detection of CIM+Carba+ isolates through targeted screening because of increased risk for CPE carriage is expected (27). Further analysis showed that the Ambler class C-type genes in our study were chromosomally encoded, and these isolates harbored limited number of plasmid replicons, suggesting that there are opportunities for these bacteria to acquire additional plasmid-based resistance mechanisms (28).

In 2019, Jousset *et al.* identified CIM+ *E. kobei* ST125 harbouring *bla*_ACT-28_, and discussed that it could be falsely classified as CPE (14), while Bonnin *et al.* described the *bla*_ACT-28_ as a minor class C carbapenemase (13). The classification of Ambler class C-type β-lactamase genes regarding their ability to produce enzymes with carbapenemase activity varies across different databases. For instance, β-lactamase database by Naas *et al.* (http://www.bldb.eu/) does not classify ACT enzymes as carbapenemases, whereas CARD does (12). Our study suggests that *Enterobacter* spp. harboring *bla*_ACT_ and *bla*_MIR_ alleles, along with a positive CIM and CarbaNP, exhibit putative carbapenemase activity. Additional testing on *E. kobei* containing *bla*_ACT-28_, showed in an exemplary case that ACT-28 protein is overproduced in the CIM. The classification of CIM+Carba-*Enterobacter* carrying Ambler class C β-lactamases is challenging. There are currently no national guidelines for the CIM+Carba-*Enterobacter* spp. or *K. aerogenes* available. To classify CIM+Carba-*Enterobacter* spp. and *K. aerogenes* as CPE or as non-CPE leads to different clinical implications, particularly in the Dutch healthcare setting. Since 1^st^ July 2019, it is mandatory to notify CPE to Municipal Health Services whereas this is not needed for non-CPE isolates (29). In the Netherlands, healthcare professionals should take standard infection prevention precautions, such as hand hygiene and wearing appropriate clothing, when caring for patients with Enterobacterales that are non-CPE. The use of personal protective equipment (PPE) is only needed in cases of direct care contact. However, when healthcare professionals care for patients with CPE in intramural settings, they should always use PPE in addition to standard precautions. Patients with CPE should be placed and treated in a single-occupancy room, and additional disinfection measures of the room are in place (29). For the national mandatory CPE reporting, CPE is currently defined as Enterobacterales that produces carbapenemases, either phenotypically or genotypically confirmed. Enterobacterales with carbapenem-resistance caused due to another mechanism than carbapenemase production are not notifiable (29). Despite not all CPE are carbapenem-resistant, CPE may act as reservoirs for transferable resistance genes, hence public health attention on this group is important. Therefore, low screening threshold (MIC >0.25 mg/l for meropenem, and/or >1 mg/l for imipenem) for confirmation of the presence of carbapenemases is advised in national guidelines in the Netherlands (9, 30). It is therefore recommended that guidelines are developed to classify CIM+Carba-*Enterobacter* spp. and *K. aerogenes* with Ambler class C-type β-lactamases as non-CPE based on whole-genome sequencing. MALDI-TOF MS is routinely used for bacterial identification but known to be insufficient to distinguish *Enterobacter* spp. pgMLST is used for *Enterobacter* spp. transmission and outbreak investigations, however it is not able to distinguish the different species within the *E. cloacae* complex (31, this study). Using whole-genome sequencing in combination with MASH analysis enabled *Enterobacter* species determination and classification of Ambler class C-type β-lactamase genes. Some *Enterobacter* species failed to be assigned to specific genogroups and may represent potential novel species. Future studies should combine biochemical and further genomic characterization of these potential novel species.

This study has some limitations. The completeness of the Dutch national surveillance dataset is uncertain as isolates are sent to the RIVM on a voluntary basis and only when meropenem and/or imipenem MICs were elevated and carbapenemase production was suspected. However, since July 1st, 2019, it is mandatory to notify CPE. Consequently, from that date forward more insights into the completeness of the database was obtained. It is possible that asymptomatic CPE carriers not meeting criteria for isolation and testing upon hospital admission will not be identified. (29). Our study only marked the five major carbapenemase genes as Carba+, however, genes such as *bla*_FRI_, *bla*_GES_ and *bla*_IMI_ are also known for its carbapenemase activity (4), but have been marked as Carba-thus far. Secondly, in-depth analysis could only be performed using NGS data. Since NGS data was not available for all isolates, further analysis was conducted only on *Enterobacter* and *K. aerogenes* isolates for which NGS data was available. Thirdly, the selected CIM-Carba-isolates show high MICs for meropenem and were therefore sent to the national surveillance. These CIM-Carba-*Enterobacter* do not show carbapenemase production and lack carbapenemase genes, resistance for these isolates is attributed to mechanisms other than carbapenemase production and were beyond the scope of this work. A further limitation of this study is the limited and/or incomplete data on susceptibility testing for other additional carbapenem antibiotics.

## Conclusion

A large proportion of *Enterobacter* and *K. aerogenes* isolates (31.6% and 52.0%, respectively) submitted to the Dutch national CPE surveillance are associated with a positive CIM and carry Ambler class C β-lactamase genes *bla*_ACT-28_, *bla*_ACT-52_, *bla*_MIR-3_, *bla*_MIR-11_ or *ampC*(-like), in the absence of major carbapenemase genes (IMP, KPC, NDM, OXA-48-like, VIM). For the CIM+Carba-*Enterobacter/K. aerogenes* isolates analysed, the Ambler class C β-lactamases may display weak carbapenemase activity, and can be overproduced in the CIM. The genes encoding these enzymes are encoded from the chromosome, and the majority of these isolates (81.9%) are susceptible for meropenem. Nosocomial spread occurred, but on a very limited scale. Health care professionals should be aware of CIM+Carba-*Enterobacter*/*K. aerogenes* isolates lacking major carbapenemases. Future national and international guidelines describing CPE are recommended to be amended to create more awareness of this low-risk group of microorganisms.

## Data Availability

Both raw NGS and Nanopore long-read sequence data are available at the sequence read archive (SRA; PRJEB35685, PRJNAB903550, PRJNA1076808 and PRJNA1122997).

## Acknowledgements

† In remembrance and dedicated to Maureen Ouw (1996-2024), who contributed to this study. We thank all the members of the Dutch CPE surveillance study Group and the Dutch medical microbiology laboratories for submitting Enterobacterales isolates to the RIVM for the national CPE surveillance program. Members of the Dutch CPE Surveillance Study Group:

- A.L.E. van Arkel, ADRZ medisch centrum, Department of Medical Microbiology, Goes
- M.A. Leversteijn-van Hall, Alrijne Hospital, Department of Medical Microbiology, Leiden
- W. van den Bijllaardt, Amphia Hospital, Microvida Laboratory for Microbiology, Breda
- R. van Mansfeld, Amsterdam UMC - location AMC, Department of Medical Microbiology and Infection Prevention, Amsterdam
- K. van Dijk, Amsterdam UMC - location Vumc, Department of Medical Microbiology and Infection Control, Amsterdam
- B. Zwart, Atalmedial, Department of Medical Microbiology, Amsterdam
- B.M.W. Diederen, Bravis Hospital/ZorgSaam Hospital Zeeuws-Vlaanderen, Department of Medical Microbiology, Roosendaal/Terneuzen
- H. Berkhout, Canisius Wilhelmina Hospital, Department of Medical Microbiology and Infectious Diseases, Nijmegen
- A. Ott, Certe, Department of Medical Microbiology Groningen & Drenthe, Groningen
- K. Waar, Certe, Department of Medical Microbiology Friesland & Noordoostpolder, Leeuwarden
- W. Ang, Comicro, Department of Medical Microbiology, Hoorn
- J. da Silva, Deventer Hospital, Department of Medical Microbiology, Deventer
- A.L.M. Vlek, Diakonessenhuis Utrecht, Department of Medical Microbiology and Immunology, Utrecht
- J.J.J.M. Stohr, Elisabeth-TweeSteden (ETZ) Hospital, Microvida laboratory for Microbiology, Tilburg
- L.G.M. Bode, Erasmus University Medical Center, Department of Medical Microbiology and Infectious Diseases, Rotterdam
- A. Jansz, Eurofins PAMM, Department of Medical Microbiology, Veldhoven
- S. Paltansing, Franciscus Gasthuis & Vlietland, Department of Medical Microbiology and Infection Control, Rotterdam
- A.J. van Griethuysen, Gelderse Vallei Hospital, Department of Medical Microbiology, Ede
- J.R. Lo Ten Foe, Gelre Hospital, Department of Medical Microbiology and Infection Control, Apeldoorn
- M.J.C.A. van Trijp, Groene Hart Ziekenhuis, Department of Medical Microbiology and Infection Prevention, Gouda
- M. Wong, Haga Hospital, Department of Medical Microbiology, ‘s-Gravenhage
- A.E. Muller, HMC Westeinde Hospital, Department of Medical Microbiology, ‘s-Gravenhage
- M.P.M. van der Linden, IJsselland hospital, Department of Medical Microbiology, Capelle a/d IJssel
- M. van Rijn, Ikazia Hospital, Department of Medical Microbiology, Rotterdam
- S.B. Debast, Isala Hospital, Laboratory of Medical Microbiology and Infectious Diseases, Zwolle
- E. Kolwijck, Jeroen Bosch Hospital, Department of Medical Microbiology and Infection Control, ‘s-Hertogenbosch
- N. Al Naiemi, LabMicTA, Regional Laboratory of Microbiology Twente Achterhoek, Hengelo
- T. Schulin, Laurentius Hospital, Department of Medical Microbiology, Roermond
- S. Dinant, Maasstad Hospital, Department of Medical Microbiology, Rotterdam
- S.P. van Mens, Maastricht University Medical Centre, Department of Medical Microbiology, Infectious Diseases & Infection Prevention, Maastricht
- D.C. Melles, Meander Medical Center, Department of Medical Microbiology, Amersfoort
- J.W.T. Cohen Stuart, Noordwest Ziekenhuisgroep, Department of Medical Microbiology, Alkmaar
- P. Gruteke, OLVG Lab BV, Department of Medical Microbiology, Amsterdam
- A.P. van Dam, Amsterdam Health Service, Public Health Laboratory, Amsterdam
- I. Maat, Radboud University Medical Center, Department of Medical Microbiology, Nijmegen
- B. Maraha, Regional Laboratory for Microbiology, Department of Medical Microbiology, Dordrecht
- J.C. Sinnige, Regional Laboratory of Public Health, Department of Medical Microbiology, Haarlem
- E. van der Vorm, Reinier de Graaf Groep, Department of Medical Microbiology, Delft
- M.P.A. van Meer, Rijnstate Hospital, Laboratory for Medical Microbiology and Immunology, Velp
- M. de Graaf, Saltro Diagnostic Centre, Department of Medical Microbiology, Utrecht
- E. de Jong, Slingeland Hospital, Department of Medical Microbiology, Doetinchem
- S.J. Vainio, St Antonius Hospital, Department of Medical Microbiology and Immunology, Nieuwegein
- E. Heikens, St Jansdal Hospital, Department of Medical Microbiology, Harderwijk
- M. den Reijer, Star-shl diagnostic centre, Department of Medical Microbiology, Rotterdam
- J.W. Dorigo-Zetsma, TergooiMC, Central Bacteriology and Serology Laboratory, Hilversum
- A. Troelstra, University Medical Center Utrecht, Department of Medical Microbiology, Utrecht
- E. Bathoorn, University of Groningen, Department of Medical Microbiology, Groningen
- J. de Vries, VieCuri Medical Center, Department of Medical Microbiology, Venlo
- D.W. van Dam, Zuyderland Medical Centre, Department of Medical Microbiology and Infection Control, Sittard-Geleen
- E.I.G.B. de Brauwer, Zuyderland Medical Centre, Department of Medical Microbiology and Infection Control, Heerlen
- R. Steingrover, St. Maarten Laboratory Services, Department of Medical Microbiology, Cay Hill (St. Maarten)
- Analytical Diagnostic Center N.V. Curaçao, Department of Medical Microbiology, Willemstad (Curaçao)

## Transparency declaration

The authors have nothing to disclose.

---

**Supplementary Table S1.** Genomic location of Ambler class C-type genes in *E. kobei* and *E. roggenkampii*.

| Strain | *Enterobacter*<br>spp. | Contig<br>length (bp) | GC | *bla*<sub>ACT-</sub><br>9 | *bla*<sub>ACT-</sub><br>28 | *bla*<sub>ACT-</sub><br>51 | *bla*<sub>ACT-</sub><br>52 | *bla*<sub>MIR-</sub><br>3 | *bla*<sub>MIR-</sub><br>10 | *bla*<sub>MIR-</sub><br>16 | *bla*<sub>MIR-</sub><br>22 |
| --- | --- | --- | --- | --- | --- | --- | --- | --- | --- | --- | --- |
| cRIVM_C009458_1 | *E. kobei* | 4,708,241 | 55.0% |  | + |  |  |  |  |  |  |
| cRIVM_C009974_1 | *E. kobei* | 4,731,208 | 54.9% |  | + |  |  |  |  |  |  |
| cRIVM_C011296_1 | *E. kobei* | 5,042,444 | 54.8% |  | + |  |  |  |  |  |  |
| cRIVM_C014068_1 | *E. kobei* | 4,844,744 | 54.8% |  | + |  |  |  |  |  |  |
| cRIVM_C018974_1 | *E. kobei* | 4,712,553 | 54.9% |  | + |  |  |  |  |  |  |
| cRIVM_C036326_1 | *E. kobei* | 4,855,475 | 54.9% |  | + |  |  |  |  |  |  |
| cRIVM_C038000_1 | *E. kobei* | 4,786,212 | 54.9% |  | + |  |  |  |  |  |  |
| cRIVM_C039954_1 | *E. roggenkampii* | 4,727,570 | 56.2% |  |  |  |  |  |  | + |  |
| cRIVM_C040296_1 | *E. kobei* | 4,771,999 | 55.1% |  | + |  |  |  |  |  |  |
| cRIVM_C044893_1 | *E. roggenkampii* | 4,854,898 | 56.0% |  |  |  |  | + |  |  |  |
| cRIVM_C046776_1 | *E. kobei* | 4,726,403 | 55.1% |  | + |  |  |  |  |  |  |
| cRIVM_C050138_1 | *E. roggenkampii* | 4,659,403 | 56.1% |  |  |  |  |  | + |  |  |
| cRIVM_C050142_1 | *E. kobei* | 4,875,129 | 55.0% |  |  |  | + |  |  |  |  |
| cRIVM_C055186_1 | *E. kobei* | 4,767,850 | 55.0% |  |  | + |  |  |  |  |  |
| cRIVM_C059300_1 | *E. roggenkampii* | 4,786,153 | 56.1% |  |  |  |  | + |  |  |  |
| cRIVM_C059468_1 | *E. kobei* | 4,911,345 | 54.9% | + |  |  |  |  |  |  |  |
| cRIVM_C059516_1 | *E. roggenkampii* | 4,691,741 | 55.9% |  |  |  |  | + |  |  |  |
| cRIVM_C059867_1 | *E. kobei* | 4,997,629 | 54.8% |  |  |  | + |  |  |  |  |
| cRIVM_C065837_1 | *E. roggenkampii* | 4,597,662 | 56.1% |  |  |  |  |  |  |  | + |

Supplementary Figure S1. **Determination of the MASH distance cut-off for *Enterobacter* spp./*K. aerogenes* species determination.**

Supplementary Figure S2. **Genetic clusters for CIM+Carba-*Enterobacter* isolates using pan-genome multi-locus sequence typing (pgMLST).** Five genetic clusters (≥2 isolates varying ≤20 pgMLST alleles) were observed and indicated by coloured halo.

Supplementary Figure S3. **wgMLST minimum spanning tree of CIM+Carba-*K. aerogenes* isolates.** No genetic clusters (≥2 isolates varying ≤20 wgMLST alleles) were observed.

Supplementary Figure S4. **Example of overproduction of Ambler class C-type ACT-28 enzyme of *E. kobei* with (2 isolates) or without *bla*_ACT-28_ (1 isolate) in reducing SDS-PAGE.** The ACT-28 protein sequence including signal peptide is depicted below the SDS-PAGE gel, and in red the ACT-28 polypeptides detected by Nano-LC-MS/MS.

## References

1. Tängdén T, Giske CG. 2015. Global dissemination of extensively drug-resistant carbapenemase-producing Enterobacteriaceae: clinical perspectives on detection, treatment and infection control. J Intern Med 277:501–512.

2. World Health Organization. 2024. WHO bacterial priority pathogens list, 2024: Bacterial pathogens of public health importance to guide research, development and strategies to prevent and control antimicrobial resistance. World Health Organization. https://www.who.int/publications/i/item/9789240093461.

3. Queenan AM, Bush K. 2007. Carbapenemases: The Versatile β-Lactamases. Clin Microbiol Rev 20:440–458.

4. Naas T, Oueslati S, Bonnin RA, Dabos ML, Zavala A, Dortet L, Retailleau P, Lorga BI. 2017. Beta-lactamase database (BLDB) – structure and function. J Enzyme Inhib Med Chem 32:917–919.

5. Ambler RP, Coulson AFW, Frère JM, Ghuysen JM, Joris B, Forsman M, Levesque RC, Tiraby G, Waley SG. 1991. A standard numbering scheme for the class A β-lactamases. Biochem J 276:269–270.

6. Pitout JDD, Peirano G, Kock MM, Strydom K-A, Matsumura Y. 2019. The Global Ascendency of OXA-48-Type Carbapenemases. Clin Microbiol Rev 33:e00102–19.

7. Peirano G, Pitout JDD. 2025. Rapidly spreading Enterobacterales with OXA-48-like carbapenemases. J Clin Microbiol e01515–24.

8. Davin-Regli A, Lavigne J-P, Pagès J-M. 2019. Enterobacter spp.: Update on Taxonomy, Clinical Aspects, and Emerging Antimicrobial Resistance. Clin Microbiol Rev 32:e00002–19.

9. van der Zwaluw K, Witteveen S, Wielders L, van Santen M, Landman F, de Haan A, Schouls LM. 2020. Molecular characteristics of carbapenemase-producing Enterobacterales in the Netherlands; results of the 2014–2018 national laboratory surveillance. Clin Microbiol Infect 26:1412.e7–1412.e12.

10. Wyres KL, Holt KE. 2018. Klebsiella pneumoniae as a key trafficker of drug resistance genes from environmental to clinically important bacteria. Curr Opin Microbiol 45:131–139.

11. Jacoby GA. 2009. AmpC Beta-Lactamases. Clin Microbiol Rev 22:161–182.

12. Alcock BP, Huynh W, Chalil R, Smith KW, Raphenya A, Wlodarski MA, Edalatmand A, Petkau A, Syed SA, Tsang KK, Baker SJC, Dave M, McCarthy M, Mukiri KM, Nasir JA, Golbon B, Imtiaz H, Jiang X, Kaur K, Kwong M. 2022. CARD 2023: expanded curation, support for machine learning, and resistome prediction at the Comprehensive Antibiotic Resistance Database. Nucleic Acids Res 51:D690–D699.

13. Bonnin RA, Jousset AB, Emeraud C, Oueslati S, Dortet L, Naas T. 2021. Genetic Diversity, Biochemical Properties, and Detection Methods of Minor Carbapenemases in Enterobacterales. Front Med 7:616490.

14. Jousset AB, Saoussen Oueslati, Sandrine Bernabeu, Takissian J, Elodie Creton, Vogel A, Aimie Sauvadet, Garance Cotellon, Gauthier L, Laurent Dortet, Naas T. 2019. False-Positive Carbapenem-Hydrolyzing Confirmatory Tests Due to ACT-28, a Chromosomally Encoded AmpC with Weak Carbapenemase Activity from Enterobacter kobei. Antimicrob Agents Chemother 63:e02388–18.

15. Wielders CCH, Schouls LM, H S Woudt S, Notermans DW, Hendrickx APA, Bakker J, Kuijper EJ, Schoffelen AF, de Greeff SC. 2022. Epidemiology of carbapenem-resistant and carbapenemase-producing Enterobacterales in the Netherlands 2017–2019. Antimicrob Resist Infect Control 11:57.

16. van der Zwaluw K, de Haan A, Pluister GN, Bootsma HJ, de Neeling AJ, Schouls LM. 2015. The Carbapenem Inactivation Method (CIM), a Simple and Low-Cost Alternative for the Carba NP Test to Assess Phenotypic Carbapenemase Activity in Gram-Negative Rods. PLoS ONE 10:e0123690.

17. Landman F, Jamin C, de Haan A, Witteveen S, Bos J, van der Heide HGJ, Schouls LM, Hendrickx APA. 2024. Genomic surveillance of multidrug-resistant organisms based on long-read sequencing. Genome Med 16:137.

18. Hendrickx APA, Debast S, Pérez-Vázquez M, Schoffelen AF, Notermans DW, Landman F, Wielders CCH, Cañada Garcia JE, Flipse J, de Haan A, Witteeen S, van Santen-Verheuvel M, de Greef SC, Kuijper E, Schouls LM. 2021. A genetic cluster of MDR Enterobacter cloacae complex ST78 harbouring a plasmid containing blaVIM-1 and MCR-9 in the Netherlands. JAC Antimicrob Resist 3:dlab046.

19. Hendrickx APA, Landman F, de Haan A, Borst D, Witteveen S, van Santen-Verheuvel MG, van der Heide HGJ, Schouls LM. 2020. Plasmid diversity among genetically related Klebsiella pneumoniae blaKPC-2 and blaKPC-3 isolates collected in the Dutch national surveillance. Sci Rep 10:16778.

20. Hendrickx APA, Landman F, de Haan A, Witteveen S, van Santen-Verheuvel MG, Schouls LM. 2021. BlaOXA-48-like genome architecture among carbapenemase-producing Escherichia coli and Klebsiella pneumoniae in the Netherlands. Microb Genom 7:000512.

21. Loman NJ, Quinlan AR. 2014. Poretools: a toolkit for analyzing nanopore sequence data. Bioinformatics 30:3399–3401.

22. Wick RR, Judd LM, Gorrie CL, Holt KE. 2017. Unicycler: Resolving bacterial genome assemblies from short and long sequencing reads. PLoS Comput Biol 13:e1005595.

23. Schwengers O, Jelonek L, Dieckmann MA, Beyvers S, Blom J, Goesmann A. 2021. Bakta: rapid and standardized annotation of bacterial genomes via alignment-free sequence identification. Microb Genom 7:000685.

24. Ondov BD, Treangen TJ, Melsted P, Mallonee AB, Bergman NH, Koren S, Phillippy AM. 2016. Mash: fast genome and metagenome distance estimation using MinHash. Genome Biol 17:132.

25. Shannon P. 2003. Cytoscape: a Software Environment for Integrated Models of Biomolecular Interaction Networks. Genome Res 13:2498–504.

26. Feng Y, Hu Y, Zong Z. 2021. Reexamining the Association of AmpC Variants with Enterobacter Species in the Context of Updated Taxonomy. Antimicrob Agents Chemother 65:e0159621.

27. Glasner C, Albiger B, Buist G, Tambić Andrašević A, Cantón R, Carmeli Y, Friedrich AW, Giske CG, Glupczynski Y, Gniadkowski M, Livermore DM, Nordmann P, Poirel L, Rossolini GM, Seifert H, Vatopoulos A, Walsh T, Woodford N, Donker T, Monnet DL. 2013. Carbapenemase-producing Enterobacteriaceae in Europe: a survey among national experts from 39 countries, February 2013. Eurosurveillance 18:20525.

28. Rodríguez-Beltrán J, DelaFuente J, León-Sampedro R, MacLean RC, San Millán Á. 2021. Beyond horizontal gene transfer: the role of plasmids in bacterial evolution. Nat Rev Microbiol 19:1–13.

29. Nederlandse Vereniging voor Medische Microbiologie. 2024. Bijzonder resistente micro-organismen (BRMO) | SRI-richtlijnen. https://www.sri-richtlijnen.nl/brmo. Retrieved 8 February 2025.

30. Nederlandse Vereniging voor Medische Microbiologie. 2021. Richtlijn Laboratoriumdetectie bijzonder resistente micro-organismen (BRMO). https://richtlijnendatabase.nl/richtlijn/laboratoriumdetectie_bijzonder_resistente_micro-organismen_brmo/startpagina_-_laboratoriumdetectie_bijzonder_resistente_micro-organismen_brmo.html. Retrieved 8 February 2025.

31. Paauw A, Caspers MPM, Schuren FHJ, Leverstein-van Hall MA, Delétoile A, Montijn RC, Verhoef J, Fluit AC. 2008. Genomic Diversity within the Enterobacter cloacae Complex. PLoS ONE 3:e3018.

