## Supplementary figures and images for "Ambler class C-type β-lactamases in *Enterobacter* spp. and *Klebsiella aerogenes* in the Netherlands, 2012-2023"

### Supplementary Figure 1

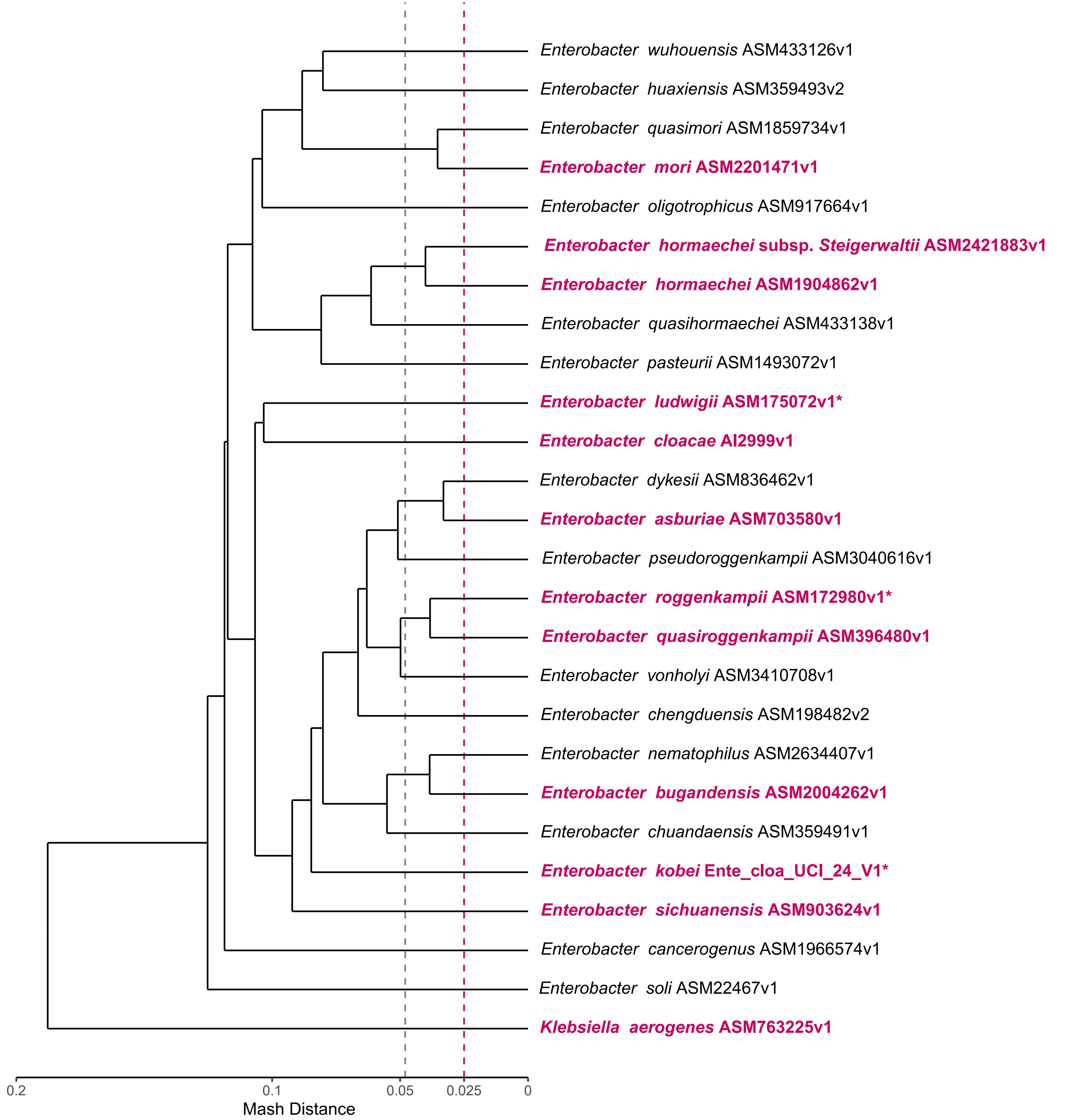

### Supplementary Figure 2

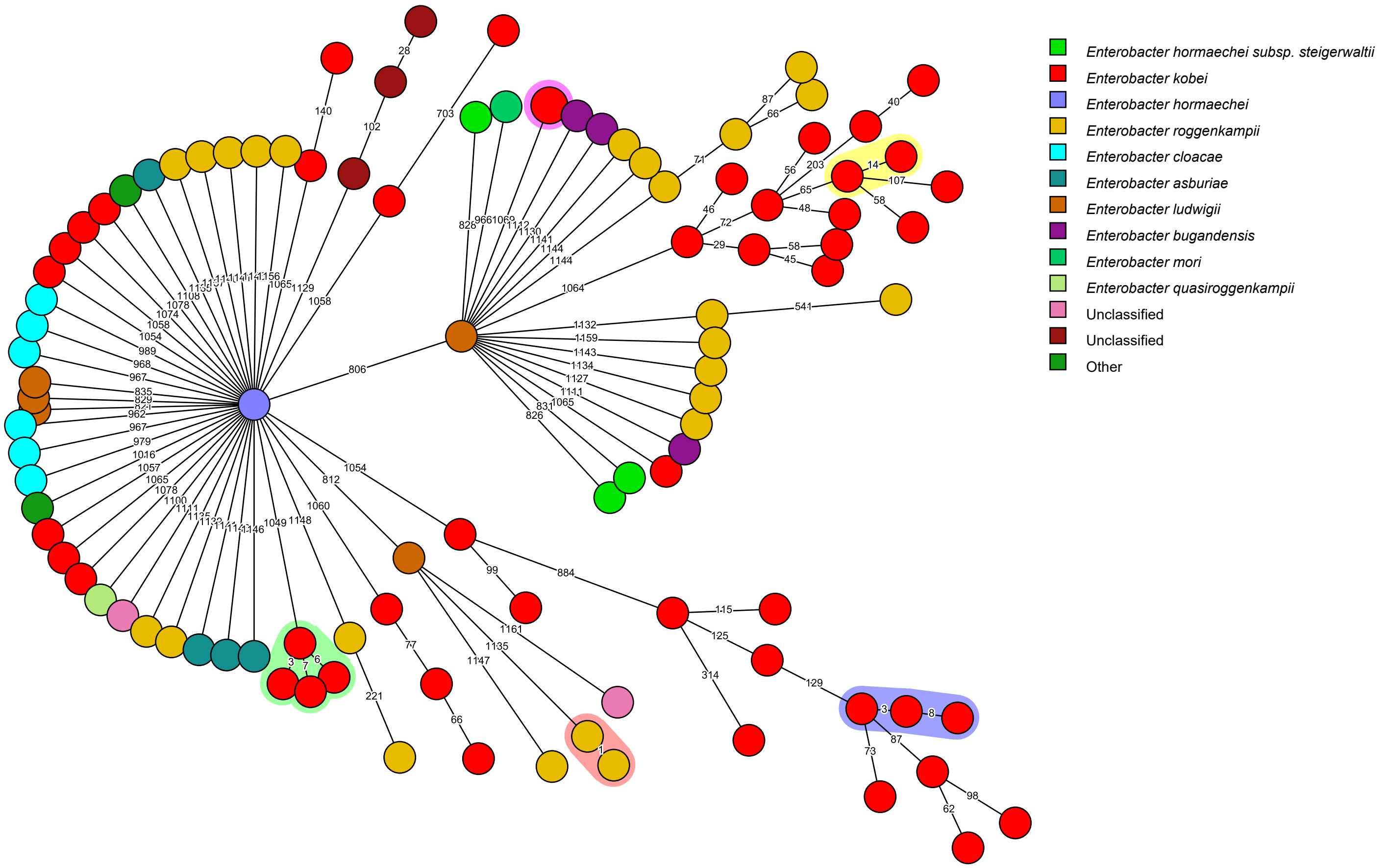

### Supplementary Figure 3

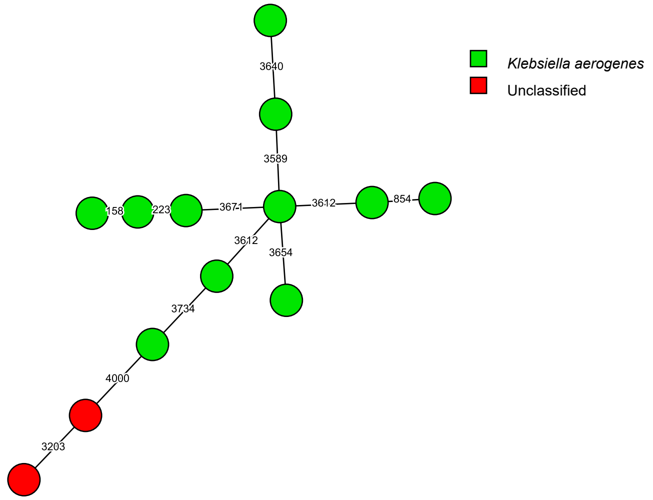
