## Supplementary Figure 4 for "Ambler class C-type β-lactamases in *Enterobacter* spp. and *Klebsiella aerogenes* in the Netherlands, 2012-2023"

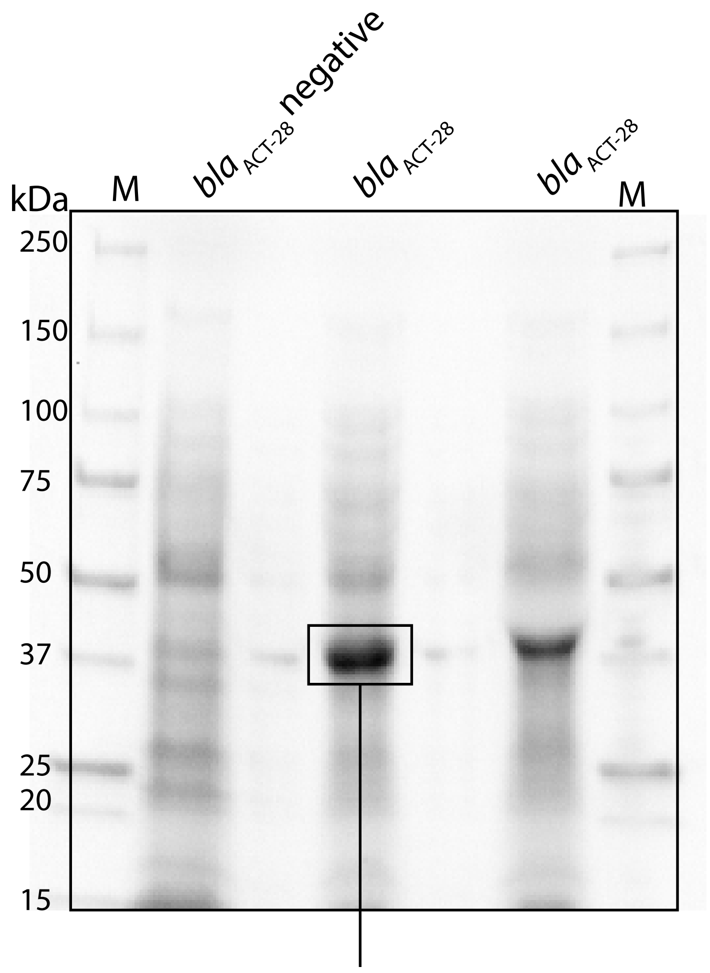


Signal peptide

```
1   MKTKSLCCAL LLSTSCSVLA |APMSEKQLSD VVERTVTPLM KAQAIPGMAV AVIYQGQPHY
61  FTFGKADVAA NKPVTPQTLF ELGSISKTFT GVLGGDAIAR GEISLGDPVT KYWPELTGKQ
121 WQGIRMLDLA TYTAGGLPLQ VPDEVTDNAS LLRFYQHWQP QWKPGATRLY ANASIGLFGA
181 LAVKPSGMSF EQAMTKRVFK PLKLDHTWIN VPKEEEAHYA WGYRDGKAIH VSPGMLDAEA
241 YGVKTNIQDM ASWLKANMNP DALPDSTLKQ GIALAQSRYW RVGAMYQGLG WEMLNWPVEA
301 KTVVEGSDNK VALAPLPVAE VNPPAPPVKA SWVHKTGSTG GFGSYVAFIP EKELGIVMLA
361 NKSYPNPARV EAAYRILSAL Q
```
